# The Cost Outcome Pathway Framework: Integrating socio-economic impacts to Adverse Outcome Pathways

**DOI:** 10.1101/2024.02.20.24303098

**Authors:** Thibaut Coustillet, Xavier Coumoul, Anne-Sophie Villégier, Michèle Bisson, Ellen Fritsche, Jean-Marc Brignon, Florence Zeman, Karine Audouze

## Abstract

Several chemical’s families are linked to a loss of intellectual quotient (IQ) points in children. This may lead to reduced working productivity and/or lower wages in adulthood and contribute to increasing the substantial socio-economic burden worldwide. The Adverse Outcome Pathway (AOP) concept, that leverages existing data to formalize knowledge, is a well-accepted concept in risk assessment although it does not handle the socio-economic impact that environment-induced diseases may generate. Here, we propose to extend the AOP framework by bridging an adverse outcome (AO) to a cost outcome (CO) creating so-called Cost Outcome Pathways (COPs) for including the socio-economic costs of exposure to chemicals. As a case study, a COP related to neurodevelopmental toxicity was designed, with a connection between the AO ‘decreased, IQ’ and the CO ‘increased, socio-economic burden’. For support to policymaking in the public health sector, this framework might also hold great potential for environmental exposure-related diseases such as cancer or obesity which are diseases with known detrimental socio-economic impacts.

## Introduction

The exposome concept, proposed by C.P. Wild in 2005, gathers all exposure sources (environmental chemicals, biological agents, physical & socio-economic factors…) to which an individual is subjected, from conception to death^1^. Complementary to the genome approach, the exposome is an important notion for the understanding of how gene-environment interactions trigger diseases^2^. The environmental contribution to the development of chronic diseases could be in the range of 70% to 90%^3^. Agrochemicals that include biocides, herbicides, fungicides, and insecticides contribute chronically and significantly to the chemical exposome worldwide due to their extensive use in agriculture and domestic environments. In Europe, different families of man-made pesticides have been used in waves since the 1950s, notably in agriculture. For example, the organochlorines (e.g., DDT) were replaced by organophosphates (OPs), which are now progressively replaced by pyrethroids. Despite their gradual substitutions by pyrethroids in Europe, some OPs including chlorpyrifos (CPF), are still widely used and sometimes benefit from exemptions for their use (e.g., France, spinach cultivation). Recently, the National Institute of Health and Medical Research in France (INSERM) published a collective expert report identifying strong presumed risks associated with exposure to OPs^4^ (non-Hodgkin’s lymphomas or cognitive disorders in adults and altered motor, cognitive and sensory capacities in children due to exposure during pregnancy).

Although exposure to risk factors increases the probability of acquiring an adverse outcome, the window, timespan, and dose of exposure are elements to be considered when assessing the hazardousness of a chemical. In line with the Developmental Origins of Health and Disease (DOHaD) theory, which emphasizes linkage between prenatal and postnatal exposure to environmental factors and the risk of developing several diseases later in life^5^, the perinatal period is a critical window of high vulnerability for neurodevelopment^6^. Therefore, children’s exposure to environmental pollutants (through contaminated food/air/water, direct contact through crop spraying, domestic use, etc.), may result in behavioral and/or cognitive disorders, potentially persistent through adult life^7^.

To have a better understanding of the putative involvement from the chemical exposome to health effects, new and innovative methods are needed. New Approaches Methodologies (NAMs) or next generation risk assessment (NGRA), referring to non-animal-based approaches, can provide information on chemical hazards and inform risk assessment^8^. In 2010, Ankley *et al*. formalized the Adverse Outcome Pathway (AOP) framework^9^ which compile existing biological knowledge in a structured linear representation at various levels of the biological organization (molecular, cellular, individual, etc.). An AOP starts with a Molecular Initiating Event (MIE), induced by stressor(s), that lead to an Adverse Outcome (AO) *via* a finite number of Key Events (KEs), where each KE is connected to its neighbor by a Key Event Relationship (KER). Although stressors (i.e., the exposome), trigger the MIE and thus the entire downstream AOP, they are not part of AOPs due to the stressor-agnostic nature of them. AOPs are thought to provide a mechanistic framework for interpretation of results produced with NAMs and conventional studies^10^. Advancements in technologies, coupled with an uptick in data volume and the availability of diverse toxicological data from various sources (omics, high throughput assays, literature), facilitate the development of complex, but realistic toxicological models including AOPs^11^. As the AOP construction could be very time-intensive to compile existing heterogeneous knowledge from structured- and non-structured data, innovative computational methods based on artificial intelligence (AI), and data mining technologies are well suited. AI allows to identify, extract, and compile relevant sparse information from the wealth of available open-source data, and can be used for predictive toxicology (e.g., Abstract Sifter^12^, the ComptoxAI tools^13,14^) or others computational approaches^15^. Recently, AI and text-mining were used to develop AOP-helpFinder^16,17^, a tool to automatically identify, extract, and prioritize knowledge from the literature (PubMed) (https://aop-helpfinder.u-paris-sciences.fr/)^18^. AOP-helpFinder has already been successfully used in several studies^19–23^, as well as in the development of novel AOPs that were submitted to the AOP-Wiki database (e.g., AOP IDs 439, 441, 490, 493, 494 and 497).

The AOP concept can be extended to the socio-economic consequences of AOs (or diseases) beyond its biological information (MIE and KE). Diseases may cause suffering, moral distress, loss of earnings, and set limits in the present and future life of affected individuals in addition to decrease their quality of life, as many dimensions that are not captured by the depiction of the biological KEs in AOP. Due to these consequences, and the need for healthcare and other social cares (psychological support, economic support,…), AOs may imply costs to the society that need to be reflected in AOPs as relevant consequences from chemical exposure. Notwithstanding the fact that there are studies that have already outlined, e.g., disability-adjusted life years due to diseases^24^, the socio-economic costs of disease need to be more often considered and formalized, particularly in the case of exposome-induced diseases. Here we proposed a new concept, the Cost Outcome Pathway (COP) that integrates the socio-economical cost (Cost Outcome, CO) into the AOP framework. As a case study, one AOP related to agrochemicals exposure (OPs including CPF) during neurodevelopment was designed using the AOP-helpFinder tool and scientific expertise’s. The proposed AOP was extended to a COP, by adding a ‘socio-economic KER’ describing socio-economic consequence.

## Results

### Conventional stressor-event linkage retrieved by literature screening using artificial intelligence

Out of the 24 unique chemicals and 253 events (MIEs, KEs, AOs, COs) selected as input for AOP-helpFinder 2.0, a total of 2404 stressor-event links were identified in the PubMed database (i.e., between 23 unique chemicals and 141 unique events) (see Supplementary Table 1). No association was found for the pyrethroid metabolite of tefluthrin (CICF3CA). The chemical with the most information in the PubMed database was the chlorpyrifos (representing a total of 33% of the identified knowledge), followed by deltamethrin, permethrin, cypermethrin and chlorpyrifos-oxon. Among these 2404 stressor-event linkage identified in 1507 unique scientific abstracts, 551 were unique. Two biological events (i.e., ‘*AchE inhibition*’ and ‘*deficit voltage-gated sodium channel*’) appear more studied than the others as they were retrieved in 850 publications, representing 35% of the compiled information. Among the 141 retrieved events co-mentioned with the stressors, a total of 20 events appears to be much studied as they represent 74,4% of the collected information. The distribution of the knowledge was largely skewed for both, stressors, and events, with most data relating only a subset of them (see Supplementary Fig. 1 & 2).

Figure 1 shows that there are clearly two separate mechanisms of action for OPs (AChE inhibition; 1^st^ and 4^th^ ranks) and pyrethroids (deficit of voltage-gated sodium channels; 2^nd^, 3^rd^, 7^th^, 9^th^, 10^th^; which is more precisely a prolonged opening). It seems that such links are well characterized in the literature, as AOP-helpFinder scored each association as ‘Very High’.

**Figure 1.**
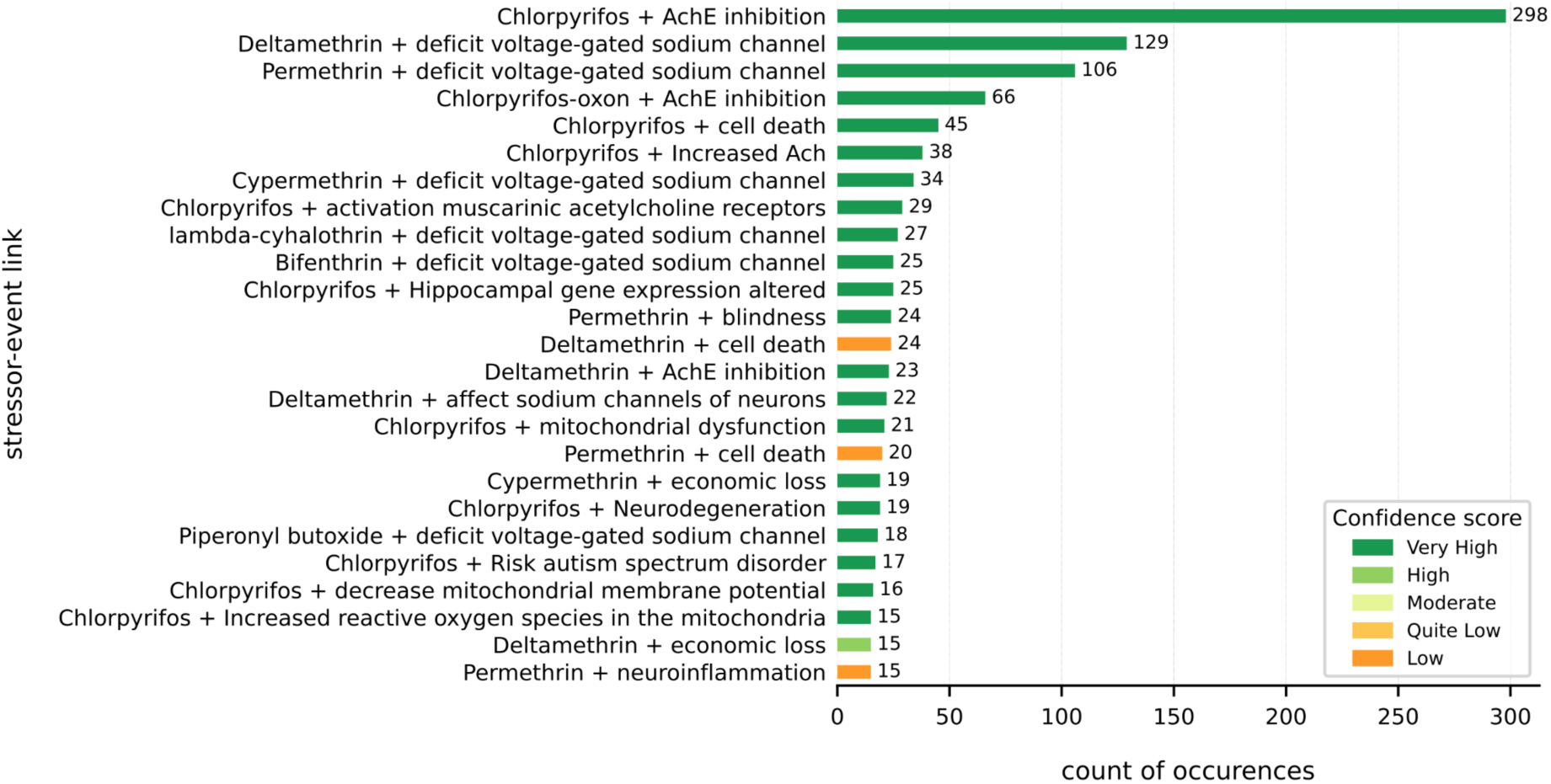
Top 25 of the most common links (stressor-event) resulting from the AOP-helpFinder v2 algorithm. The plotted distribution represents 45.3% of the total AI-obtained data set (which contains 551 different associations and 2404 total links). The color bars are based on the confidence score (CS). Numbers at the end of each row embody the numbers of PubMed abstracts that co-mention at least one stressor and one event. Results displayed here are the raw output of AOP-helpFinder 2.0, i.e. before manual curation. For convenience, only the top 25 associations were shown. The full distribution of stressor-event links can be found in the Supplementary Table 1 (see Supplemental Material). AChE, Acetylcholinesterase; ACh, Acetylcholine.

Given the extensive existing literature on CPF and AChE inhibition (Fig. 1), along with the prevalence of AOPs referencing the ‘*AchE inhibition*’ event (MIE ID12 in AOP-Wiki) and AOs related to AchE-dependent cognitive disorders (in AOP-Wiki: AOP IDs 281, 405, 450), we prioritized to introduce innovation on the AOP framework regarding CPF exposure. Even if CPF does not directly belong to the AOP, and therefore to the COP, four links between CPF and the event ‘*increased glutamate*’ (moderate CS) were found (see Supplementary Table 1).

As of today, this mechanism remains unclear, and we manually investigated a putative origin of the increase in glutamate after CPF exposure. Based on the literature, we found that CPF could trigger the release of calcium via combined activation of ryanodine receptors (RyR) and inositol trisphosphate receptors (IP3R)^25^, which could then lead to the exocytosis of glutamate from synaptic vesicles. Consequently, the ‘*Activation, IP3R*’ (ID 2037) and ‘*Activation, RyR*’ (ID 2186) events were both designated as MIEs.

### Conventional event-event linkage identified by literature screening using artificial intelligence

Regarding the event-event relationships, the AOP-helpFinder tool retrieved 73 079 links, of which 2584 were unique. Among the 253 selected events for the searches, 225 were identified. Regarding at the full distribution, only 14 events accounted for 50.5% of all obtained data (see Supplementary Fig. 3), with a very high characterization for the ‘*cell death*’ event (i.e. 12 085 publications, representing 8,3% of the extracted knowledge).17 associations were found between KEs ‘*Activation of RyR*’ and ‘*Activation of IP3R*’ (Very High CS), suggesting that both receptors are simultaneously active, and supporting the COP’s first assertion that CPF can act on both. After MIEs have been triggered, we found that both IP3R and RyR induce cell death *via* increased glutamate. Cell death involves classic protagonists with well-documented evidence, such as the Bax and Bcl-2 proteins^26^, but also the participation of microglia cells (inflammation) in the case of the nervous system^27^. Upon the occurrence of cell death in the nervous system, particularly in the hippocampus, learning and memory disorders may occur^28^ (for detailed toxicity pathway see the Supplementary Material). Regarding the full distribution of event-event associations (before manual curation), all results are available on Supplementary Table 2 (see Supplemental Material).

### Design of a conventional Adverse Outcome Pathway using intelligence artificial and expert knowledge

From this, we proposed for the first-time linkage between the biological events and the event ‘*lower IQ’*. The event ‘*deficits Working Memory Index*’ was paired 10 times with it (Very High CS), highlighting the relevance of IQ as a marker of cognitive impairment that may result from neurodevelopmental concerns related to learning and memory. Nowadays, most of the tests investigating a child’s IQ are carried out on the Wechsler Intelligence Scale for Children (WISC). Using the 4^th^ edition (WISC-IV, 2003), Rauh *et al.*, showed that for each 4.61 pg.g^-1^ increase in CPF prenatal exposure, full-scale IQ declined on average by 1.4% and working memory index scores by 2.8% in children aged 7 years^29^. Furthermore, children with lower hippocampal volume due to congenital heart disease (aged 9-11) made more mistakes in the digit span test, which consists of repeating numbers in the same order as they are presented, thereby reflecting memory dysfunctions. Although a lower hippocampal volume was not correlated to other impairments in memory tests (e.g., attentional performance), authors showed a significant association with a lower IQ^30^. On the other hand, the relationship between learning and IQ is intertwined, and therefore it is unclear to discern whether one is the cause or the consequence of the other. In children with smaller brain volume due to preterm birth, a correlation has been found between poorer performances in mathematical calculations and lower IQ^31^. A deficit in the learning of fundamentals such as mathematics may lead to poorer educational performance, which may lead to a lower IQ as learning and school results are closely intermingled. However, it is difficult to establish an oriented linear link between mathematics, learning and IQ (see KER Known Modulating Factors below). In another study, children born to mothers from poor-quality home environments have lower cognitive scores at age 7, including working memory and full-scale IQ^32^. These results suggest that IQ is highly dependent on memory and learning abilities, although the direction of causality remains a trifle unclear.

### Development of the Cost Outcome Pathway: identifying socio-economic costs resulting from the adverse outcomes by literature screening using artificial intelligence

AOP-helpFinder emphasized the connection between the event ‘*loss of productivity*’ and the three ones ‘*economic loss*’ (n= 369, Very High CS), ‘*Economic Burden*’ (n=175, Low CS) and ‘*Economic costs*’ (n=93, Low CS), underscoring here how the working productivity is strongly associated with the economic burden. The CPF-economic-loss association accounted for 10 links, with a very high CS (see Supplementary Table 1). The event ‘*economic loss*’ was ranked 7^th^ for all stressors combined, with 68 occurrences (see Supplementary Fig. 2). As AOP-helpFinder is a text-mining based algorithm, we had to be careful to distinguish between economic losses due to poor harvests (crop failure due to pests) and economic losses due to human adverse outcomes (what we are interested in here).

To be consistent with what’s already described in the literature, and to stay within the conventionally used terminologies, we proposed to set the CO to ‘*increased, socio-economic burden’*. The resulting COP withheld after analysis of the AOP-helpFinder results and manual curation of experts is represented in Fig 2. It was submitted as a novel AOP to the AOP-Wiki database (AOP ID 490, https://aopwiki.org/aops/490), where the event ‘*increased, socio-economic burden’* was considered as a special AO. Such a culminating socio-economic effect derives from the reduction in IQ induced by memory and learning disorders, themselves caused by the toxicity pathway described in the previous section. Indeed, in Europe, OPs (including CPF) were identified in one study as potentially responsible for the loss of 13 million IQ points in children each year when exposed prenatally^33^, contributing to an economic loss estimated as of $146 billion per year^34^, making them the stressors among the several chemicals of that study with one of the greatest impacts.

**Figure 2.**
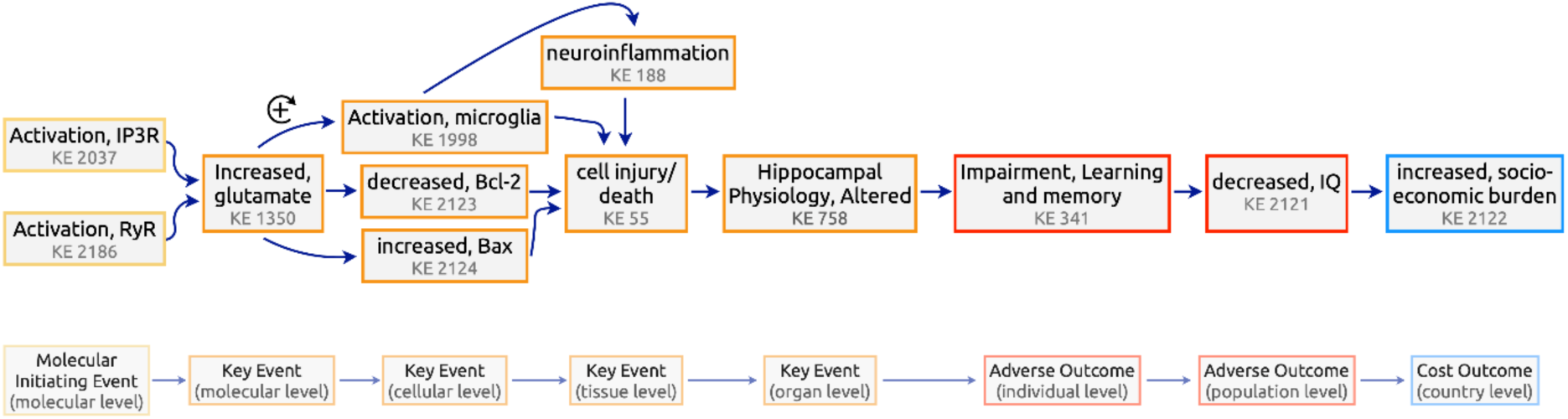
Cost Outcome Pathway. The toxicity pathway from ‘*Activation, RyR*’ and ‘*Activation, IP3R*’ to ‘*decreased, IQ*’ is the conventional AOP in the original understanding. The integration of the socio-economic event at country level expands the framework of AOP to COP by appending a CO. The plus sign surrounded by a circle represents an amplification feedback loop. Arrows indicate the Key Event Relationships (KERs). IP3R, inositol trisphosphate receptor; RyR, ryanodine receptor; IQ, intellectual quotient.

### Setting up the adverse-outcome-cost-outcome interaction

The observed association between exposure to chemicals and IQ points losses has been a motivation to understand and quantify the socio-economic implications of IQ impairment, especially in children. This COP is focusing on the direct pathway from neurological effects of agrochemicals to IQ and educational achievements, but it should be noted that similar pathways from neurotoxicological effects to other health endpoints such as for instance Attention-Deficit Hyperactivity Disorder (ADHD) or autism, and then IQ and educational achievements are also likely (Fig. 3). Also, other forms of intelligence and intellectual ability than that measured through IQ can also be affected, and therefore the social cost of agrochemicals exposure can be higher than that assessed through IQ only^35^.

**Figure 3:**
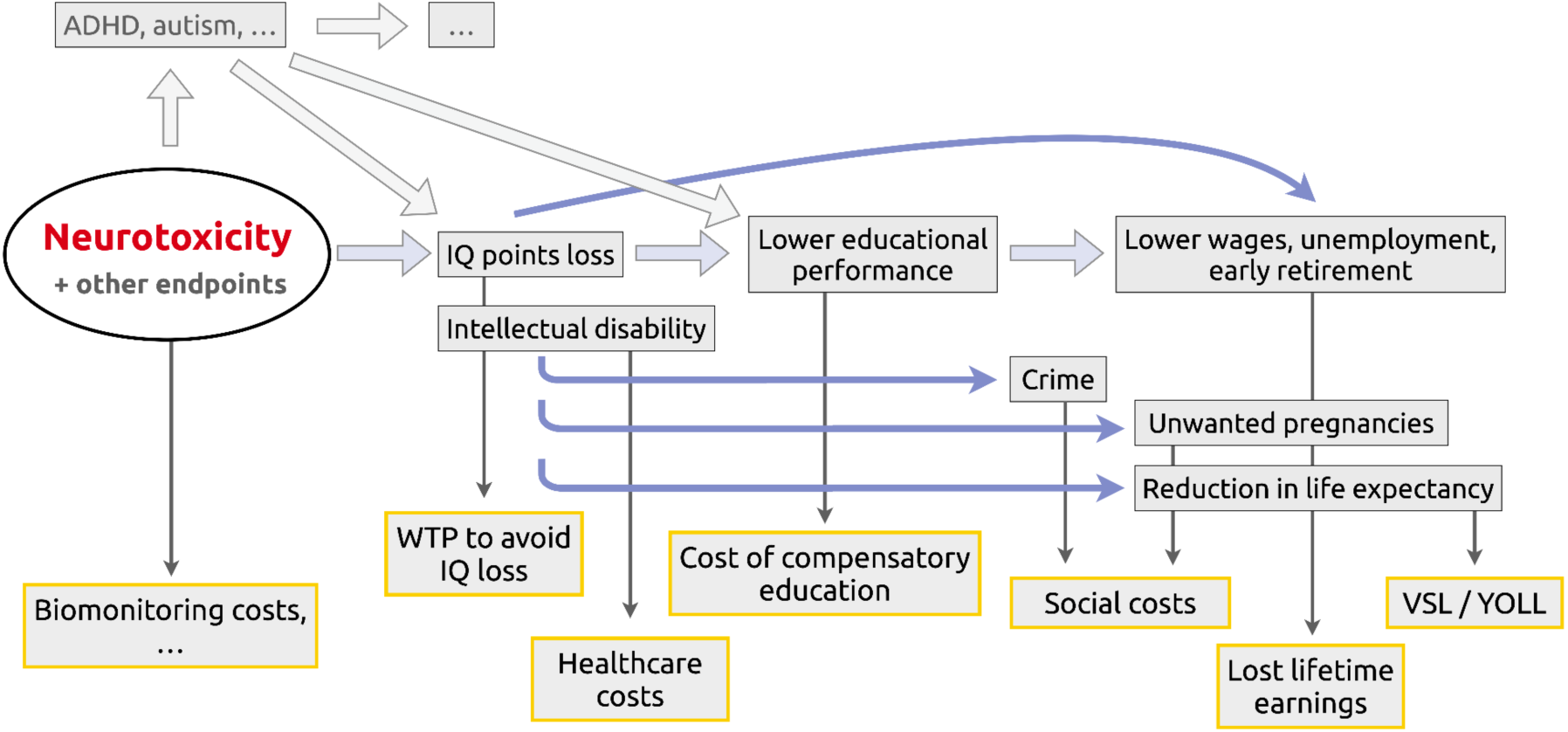
Socio-Economic consequences of IQ loss. Several paths lead to economic costs after loss of IQ points. ADHD, Attention-Deficit Hyperactivity Disorder; IQ, Intellectual Quotient; WTP, Willingness-to-pay; VSL, Value of a Statistical Life; YOLL, Year Of Life Lost.

Most studies have estimated these impacts in terms of lost discounted lifetime earnings, building on statistical association found between lower educational attainment and revenues, and between IQ and educational achievement. Lower IQ has also been found to be associated with violent behavior and crime^36,37^ and unwanted pregnancies (Fig. 3). Discounted lifetime earnings may include, depending on authors, only the impacts from lower-paid jobs or also the higher probability of being unemployed or the need to retire earlier. In 2009, E. Gould updated previous publications and took account of both the effects on earnings and employment^38^, and on crime, and found a cost per IQ point and child of €_2009_ 16 885. More recently Lin *et al.* found a value equivalent to €9 500 to €13 600 in €_2020_^39^. In 2011, C. Pichery had similarly to (Gould, 2009) included a valuation of criminal behavior, including direct costs to victims, overhead costs of justice and incarceration and lost earnings for both criminals and victims^40^. Is it of course difficult to know to which extent such associations can be extrapolated between countries and are stable over long time periods, because of the impact of socio-economic and cultural differences, and of their evolution over time.

Another approach is the human capital approach, in which the socio-economic impact of IQ loss is calculated in terms of the lost lifetime economic output of a hypothetical individual, and in practice the main difference with the lost earnings approach is that full gross salary and other indirect costs, and sometimes non-market services (like household services for non-working households), are also included^35^. Further to this, the share of the cost of compensatory education to children suffering from neurodevelopmental health impacts should be added. Data and publications are available on special education costs per child in many countries, and are likely very variable between countries with different educational systems and would also need to be apportioned to IQ loss points (if feasible), to be used in this COP. Further, since compensatory education can to some extent reduce impacts on future earnings, its cost cannot be automatically fully added to lifetime loss of revenues or to the human capital approach. Another direct and tangible economic cost of exposure to agrochemicals is the need to monitor exposure levels in the environment and in the population, to design and implement public policies, and a share of these costs could be apportioned to the individual level and to the impact of agrochemicals on IQ. Some authors make a distinction between IQ points loss (with IQ not going below 70) and mental retardation, defined as IQ passing below 70^33^. Socio-economic consequences of each case could be different, and J. Olesen *et al.* specifically valued the cost of illness of mental retardation at €_2010_ 6970 for direct costs and €_2010_ 3364 for indirect costs (but it appears that no details on the methods to fully understand difference of valuation from IQ points loss are provided in this publication)^41^.

All these tangible costs do not account for other intangible costs such as, for instance, (real and perceived) lower social achievements related to lower education and lower paid jobs, reduced overall health, aversion to IQ loss and its consequences, and shorter life expectancy that has also been associated with lower IQ. This intangible dimension can be captured when valuing IQ loss through the value placed by individuals on the reduction of the risk of losing IP points. Such surveys, that inform individuals about these consequences, have been carried out, the most recent being a Willingness-to-pay (WTP) worldwide survey by the OECD^42^, that has been focusing on presenting the consequences on educational performance, and in which the estimated mean WTP to avoid the loss of 1 IQ point was found to be USD_2022_ 609 per year and child. Reduction in life expectancy could be considered using ‘value of a statistical life (VSL)’ of ‘year of Life Lost (YOLL)’ values, which themselves also recovered from WTP or similar economic methods. It has to be stressed that tangible (based on market) and non-tangible (non-market-based) costs cannot be simply added because they are overlapping and intertwined in complex patterns^43^. Overall, in 2021 Grosse *et al.* caution that any valuation of IQ point loss is very uncertain and recommend using a value ranging from USD_2016_ 10,600 to 13,100 per child^35^.

### Robustness and overall assessment of the Cost Outcome Pathway

Results of the PubPeer test across the whole bibliography showed that none of the articles were potentially corrupt. Overall, the proposed COP is based on 34 typical scientific publications. It is not exclusively founded on this set of knowledge, as some KERs were trivial (e.g., ‘*increased, Bax leads to Cell injury/death’*) and did not necessarily need to be supported by specific publications. Regarding the PubTator bio-concepts, no trend was found. We noticed that the most frequently retrieved bio-concepts were ‘*human*’ (tagged as species; n=13), followed by ‘*glutamate*’ (tagged as chemical; n=7), ‘*neurotoxic*’ (tagged as diseases; n=6) and ‘*BCL2*’ (tagged as gene; n=5) (see Supplementary Table 3). ‘*Organophosphate*’ and ‘*chlorpyrifos*’ were among the most frequently found chemicals, supporting the relevance of the COP for this family of agrochemicals. Interestingly, the COP is particularly relevant to the neurodevelopment period as ‘*child*’ was the second most common bio-concept found in all categories (n=11).

The evidence supporting all the KERs and the ensuing COP was reviewed according to 7 features (Table 1). These criteria are a set of requirements for providing adequate evidence of a causal relationship between two KEs. The biological applicability domain of the COP focuses on human beings. Although IQ can decline at any age, we aimed to characterize the reduction during neurodevelopment. However, the consequences of a drop in IQ can be felt over the long term in adults since several socio-economic impacts of IQ loss in early life (compensatory education, reduction in earnings,…) occurs throughout life.

**Table 1:** Weight of Evidence for each KER. Each cell was filled in according to the OECD and AOP-wiki guidelines supporting the development of AOPs. Most of the criteria can be described using a three-level label: Low, Moderate or High.

| Upstream event | Downstream event | KER ID | Biological applicability | Biological Plausibility | Essentiality | Empirical Evidence | Uncertainties and | KER Known | Measurements | References |
| --- | --- | --- | --- | --- | --- | --- | --- | --- | --- | --- |

|  |  |  | ty domain | y |  |  | Inconsiste<br>ncies | Modulatin<br>g Factors | method<br>(downstre<br>am event) |  |
| --- | --- | --- | --- | --- | --- | --- | --- | --- | --- | --- |
| Activation<br>, RyR | Increased,<br>glutamate | 3036 | Mammals | Moderate | Moderate | Moderate | Low | Co-<br>activation<br>with IP3R | Microdial<br>ysis, high-<br>performan<br>ce liquid<br>chromatog<br>raphy & o-<br>phthalalde<br>hyde-<br>derived<br>fluorescen<br>ce<br>detection | Mori et<br>al., 2005 |
| Activation<br>, IP3R | Increased,<br>glutamate | 3037 | Mammals | Moderate | Moderate | Moderate | Low | Co-<br>activation<br>with RyR | Extreme<br>liquid<br>chromatog<br>raph, o-<br>phthalalde<br>hyde<br>derivatize<br>d<br>fluorescen<br>ce<br>detection | Yamamur<br>a et al.,<br>2009 |
| Increased,<br>glutamate | decreased,<br>Bcl-2<br>expression | 2868 | Mammals | High | Moderate | High | Low | - | Western<br>Blot | Nishi et<br>al., 1999<br>Phoraksa<br>et al.,<br>2023 |
| Increased,<br>glutamate | increased,<br>Bax<br>expression | 2869 | Mammals | High | Moderate | High | Low | - | Western<br>Blot | Nishi et<br>al., 1999<br>Phoraksa<br>et al.,<br>2023 |
| Increased,<br>glutamate | Activation<br>,<br>Microglia | 2861 | Mammals | Moderate | High | High | Low | - | Light<br>microscop<br>ic<br>examinati<br>on<br>Flowcyto<br>metry<br>(CD11b) | Domercq<br>et al.,<br>2013<br>Sunkaria<br>et al.,<br>2014 |
| Activation<br>,<br>Microglia | Neuroinflammation | 2862 | Mammals | High | High | High | Low | - | Sandwich<br>ELISA | Sunkaria<br>et al.,<br>2014 |
| Neuroinflammation | Cell<br>injury/death | 1687 | Mammals | High | High | High | Low | - | Trypan<br>blue<br>staining | Boje &<br>Arora,<br>1992 |
| decreased,<br>Bcl-2<br>expression | Cell<br>injury/death | 2870 | Mammals | High | Moderate | High | Low | - | Western<br>Blot of<br>cleaved<br>caspase 3,<br>Hoechst<br>33258<br>staining | Lee et al.,<br>2014 |
| increased,<br>Bax<br>expression | Cell<br>injury/death | 2871 | Mammals | High | Moderate | High | Low | - | Western<br>Blot of<br>cleaved<br>caspase 3,<br>Hoechst<br>33258<br>staining | Lee et al.,<br>2014 |
| Cell<br>injury/death | Hippocampal<br>Physiology, Altered | 2875 | rodents | High | High | High | Low | - | Hoechst<br>staining &<br>GFAP<br>staining<br>TUNEL | Wu et al.,<br>2021<br>Huang et<br>al., 2012 |
| Hippocampal<br>Physiology, Altered | Impairment, Learning<br>and<br>memory | 2864 | rodents | High | High | High | Low | - | Morris<br>water<br>maze test | Huang et<br>al., 2012<br>Su et al.,<br>2014 |
| Impairment, Learning<br>and<br>memory | decreased,<br>Intellectual Quotient | 2872 | 7-year-old<br>children | Moderate | High | Moderate | Moderate | lifestyle<br>habits | WISC-IV | Rauh et<br>al., 2011<br>Bouchard<br>et al.,<br>2011 |
| decreased,<br>Intellectual Quotient | increased,<br>Economic<br>Burden | 2867 | N/A | High | High | High | Moderate | lifestyle<br>habits | see Figure<br>3 | see 3.2.9 |

Regarding the uncertainties and inconsistencies, most KERs were labeled as ‘low’ as the scientific support was considered strong. However, the KER ID 2872 ‘*Impairment, Learning and memory leads to decreased, Intellectual Quotient*’ was considered with moderate uncertainty. In children aged 8 to 9, 3^rd^ and 4^th^ grade academic performances were both significantly influenced by IQ and study habits but not by short memory^44^. Study habits are non-cognitive variables considered to be the learning patterns that pupils set outside school, i.e. the systematic or disorganized, efficient, or non-productive way in which each student studies. As these so-called study habits can partly shape school results in the same way as IQ, we believe we can treat them as KER Known Modulating Factors, i.e. ‘*modulating factors/variables known to alter quantitative aspects of the response-response function that describes the relationship between the two KEs*’. In a wider context, we considered the personal lifestyle habits to be the KER Known Modulating Factors, i.e. habits not necessarily school dependent. By way of example, regular breakfast consumption has been correlated with a higher IQ in children^45^ while consuming screens such as television while eating has been linked to delayed language development in children^46^.

## Discussion

The present work aimed to introduce the Cost Outcome Pathway framework with a case study on assessing the hazard of CPF-exposed children during their neurodevelopment and the socio-economic consequences by using NAMs, expert knowledge and bioinformatic tools. Here, an innovative COP (AOP ID490 in the AOP-Wiki database), was designed starting with two MIEs ‘*Activation, RyR*’ and ‘*Activation, IP3R’* that lead to the KE ‘*Increased, glutamate*’, which are relevant to low doses of organophosphate CPF exposure, (i.e., CPF doses which do not necessarily inhibit AchE). For the first time, a causal association between an adverse outcome (*lower IQ*) and a social cost outcome (*increased, socio-economic burden*) was established. This led to the establishment of a new concept called Cost Outcome Pathway.

The AOP ID 490 describes effects of low-CPF-dose-induced toxicity resulting from environmental exposure. In contrast, the AOP ID 281 describes acute organophosphate poisoning (reported in suicide attempts) where glutamate release is massive. In our case study, the dose of organophosphate is below that required to inhibit AchE. The glutamate increase is thus expected to be small, but its chronicity activates microglia. As CPF exposure is found in complex mixtures in the environmental exposures, it is worthwhile to figure out the impact of other types of pesticides or factors regarding the AOP ID 490. *In vivo*, Carloni *et al.* observed about +30% glutamate in the hippocampus at middle-age rats after exposure to permethrin^47^. Although we defined the MIEs in line with the putative MoA of OP agrochemicals, the principle of COP can be relevant for additional substances such as pyrethroids. Besides, the AOP ID 490 may afford some support to previous hypotheses on neuropathology. It may support the neurochemical hypothesis for autism suggesting that it may originate from excitatory (glutamate)/inhibitory (GABA) imbalance, leading to neuronal hyper-excitability^48,49^. It may also support the therapeutic hypothesis stipulating that reduction in microglial activation may afford neuroprotection in neurodegenerative diseases^50^.

We may expect that the occurrence of the AOP ID 490 in the developing human brain will be subjected to individual differences. Numerous factors susceptible to influence the occurrence of AOP ID 490 can be listed: the window of exposure; the sex; the cocktail effect between several factors activating interspersed AOPs; environmental temperature; individual physiology (the level of metabolism). Moreover, the results of an IQ test may vary depending on several factors, including the individual’s level of fatigue, concentration, anxiety, and the quality of the relationship with the practitioner. Here, we highlighted a modulation factor for the KER ‘*Impairment, Learning and memory leads to decreased, Intellectual Quotient’* by pointing out that children’s personal habits could affect their IQ. Apical tests, such as the Total IQ test, are often preferred because they integrate the measurement of multiple functions^51^.

This COP developed at an individual level, could be used at society level to assess chemicals risk reduction policies, and maybe linked to kinetics and toxicodynamic models such as quantitative AOP. At the EU level, consideration, and evaluation of IQ points loss due to exposure to chemicals has been used in policymaking in the framework of the EU REACH regulation, to help decide on market restrictions for lead in ammunition^52^ and lead in PVC^53^. Another example of use in policymaking is in France to justify new monitoring and intervention thresholds for lead blood levels^54^. This use in policy making relies on the addition of individual effects over the population affected by that policy. It is of note that there could be, in the case of IQ, economic systemic effects since a collective decrease in cognitive capacities could have long-term and far-reaching implications in terms of functioning of the society in many areas (education, innovation and research,…). Another limitation of this individual-based approach is that it does not account for the fact that the impact of IQ loss of an individual can be ‘socio-economically transmitted’, meaning by this that lower educational and social achievement, or the consequences of crime for both victims and criminals can impair the well-being of the next generation.

The ongoing interest for AOPs is still growing in recent years. The scientific community is making a collaborative endeavor to enhance existing AOPs and propose new ones. The AOP framework is constantly evolving, and new proposals are put forward thanks to the crowdsourced collaboration. Six years after the original concept was proposed, J. Teeguarden *et al.*, proposed the notion of Aggregate Exposure Pathway (AEP) to ramify AOPs by setting out an aggregation pathway for a chemical^55^. In this case, the pathway illustrates the route taken by the chemical since its original use to the triggering of the MIE of the AOP. It is then possible to design new structures called AEP-AOP^56^. The main innovation in our study is the evolution from AOP towards COP by adding a socio-economic term at the end of the toxicity pathway. On the one hand, the study demonstrated the scalability of the AOP-helpFinder tool, initially created in relation to conventional AOP events (MIEs, KEs, AOs). The use of such AI-based tools could be applied to any type of information (e.g., COs) and no longer be restricted to the PubMed database in such a way that text-mining and artificial intelligence technologies open up new horizons for massive data mining. On the other hand, the COP framework can be applied to any AOP if there is a connection between an adverse effect and a socio-economic cost. It then reflects the economic burden borne by individuals and the care society has to take care of, that is the consequence of a biological endpoint. The last adverse effect making the AO-CO connection can be different from the decrease of IQ, as we have seen that also ADHD or autism are both associated with neurotoxicological effects and are socio-economic relevance. More generally, COPs have a wider potential interest because several pathologies associated with chemical exposure can lead to significant economic costs for society, such as cancer or obesity^57,58^, and many others. Recently, an excellent example has been the covid-19 pandemic which was subject to a colossal number of studies, some of which assessing the cost-of-illness of the disease^59^.

If AOP and diseases were more systematically extended as COPs, this would allow to describe and have a common understanding of the complex interconnections between exposures, socio-economic consequences. As Fig. 3 shows, integrating the socio-economic consequences helps reveal the interconnection between different exposure pathways and AOPs, in our case between different neurotoxicological impacts of environmental exposures. Therefore, COPs appear as a powerful tool to represent the exposome in a more holistic and consistent way, with potential to better design wide-ranging policy interventions to reduce the health and socio-economic impacts of environmental exposure to chemicals.

## Methods

To implement the new concept of COP, a multi-steps procedure has been created as illustrated on Fig. 4. The proposed approach was assessed to the putative economic impacts that may be linked to neurodevelopment.

**Figure 4.**
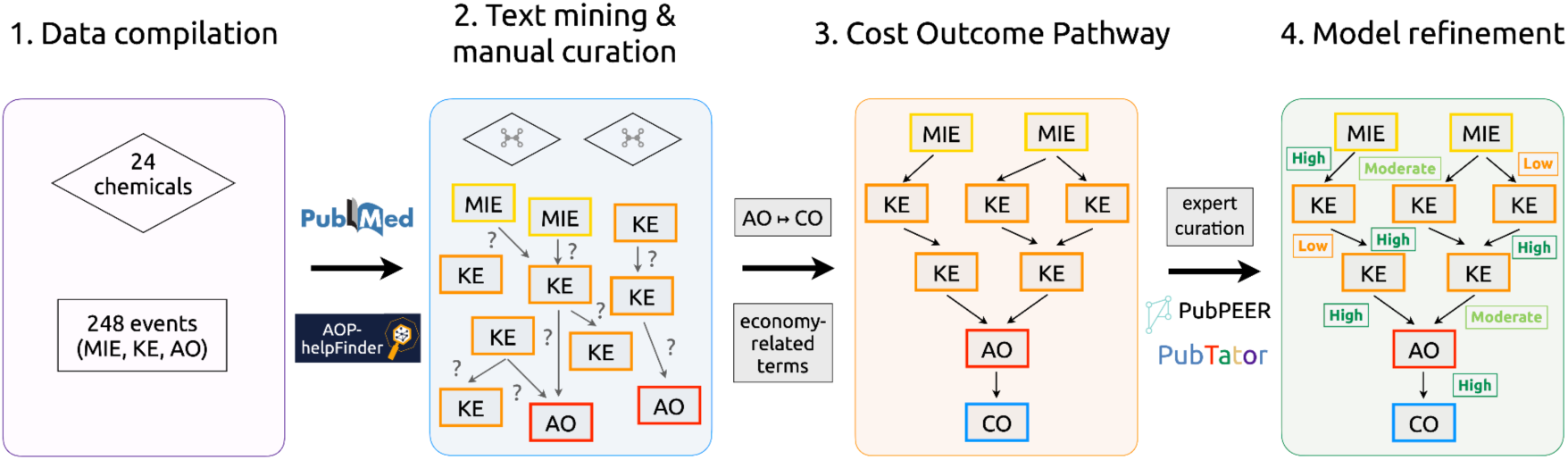
Workflow of the strategy used for building the Cost Outcome Pathway (COP). 1. Data compilation: list of chemicals and biological events defined by experts; 2. Text-mining & manual curation: the AOP-helpFinder 2.0 tool was used to automatically screen the literature (PubMed) and prioritize linkages. Then, manual curation was performed to keep the most relevant links; 3. Development of the COP: from a list of economy-related events defined by experts, the AOP-helpFinder tool was launched to decipher putative associations between Adverse Outcome (AO) and Cost Outcome (CO); 4. Model refinement: manual curation to assess the relevance of the created COP. For convenience, the COP shown here is an illustration and does not reflect the exact shape of the final COP presented in the Results section. MIE, Molecular Initiating Event; KE, Key Event; AO, Adverse Outcome; CO, Cost Outcome.

### Data compilation

#### Chemicals

Agrochemicals, widely used in France, were extracted from the National Bank of Plant Protection Products Sales by Authorized Distributors (BnVD database; https://ventes-produits-phytopharmaceutiques.eaufrance.fr/), which collects the most sold agrochemicals. Other chemical stressors were also considered based on measurements from samples (urine and hair) from the Elfe and the Esteban cohorts^60–62^, pesticides known for food contamination in France^63^, and other used for domestic applications^64,65^. Piperonyl-butoxide was also added although it is a synergist of pesticides including pyrethroids rather than a true pesticide. A synergist enhances the potency of pesticides and, in the case of piperonyl-butoxide, by inhibiting the natural detoxifying mechanisms of insects (e.g., cytochromes p450). A total of 24 unique stressors were retained for the proposed study, that includes 16 pyrethroids, 7 organophosphates, and the synergist chemical (see Supplementary Table 4).

#### Biological events

To develop an AOP linked to neurodevelopment, a list of 248 conventional events (MIEs, KEs and AOs) that are known to be involved in neurotoxicity or related biological pathways was established (see Supplementary Table 5). To be as extensive as possible, knowledge from scientific experts, usual terms from the literature, and current information from the AOP-Wiki database (as of September 2023) were compiled.

#### Socio-Economic information

To associate COs to the previously selected AOs, five CO-related terms were considered: ‘*economic costs*’, ‘*economic burden*’, ‘*economic loss*’, ‘*diminished economic productivity*’, and ‘*loss of productivity*’. These terms were screened in the literature using the AOP-helpFinder tool. As it is a text-mining algorithm based on natural language processing (NLP), introducing socio-economic-related terms did not raise any difficulties.

### Deciphering chemical-event, event-event and adverse outcome-cost outcome links by literature screening using artificial intelligence

First, to identify linkage between the 24 selected chemicals and the 248 events (MIE, KE and AO), and between the events themselves, the AOP-helpFinder 2.0 tool was ran on the PubMed database, that contains more than 36 millions of scientific articles (https://pubmed.ncbi.nlm.nih.gov/). As results, only the relevant abstracts mentioning co-occurrences, i.e. when a chemical or an event is associated with another event were provided. The selection was based on the two scoring systems proposed by AOP-helpFinder^16^. All retained abstracts were prioritized with a confidence score (CS) that statistically measure the strength of the findings (chemical-event and KER), therefore supporting the concept of biological weight of evidence (WoE). In collaboration with scientific experts, and upon these results, an AOP was suggested, pertaining to a specific AO in the context of neurodevelopment.

A second run with the AOP-helpFinder 2.0 tool was performed using the five selected COs to identify putative knowledge in the literature, to tie them to the AO present in the proposed AOP.

All analyses were performed on September 18, 2023 (date of access for the PubMed database). Regarding the AOP-helpFinder 2.0 options, the refinement filter was not used, and the threshold for skipping the introduction was set to 0%, (i.e., text mining searches were performed in full abstracts) as the aim was to collect as much information as possible.

### Development of the Cost Outcome Pathway

Although the COP is stressor-agnostic, it may be linked to some exposome-related factors to trigger the toxicity pathway. Here, as a case study, the exposure was linked to OPs, including CPF, to pinpoint the MIE in the toxicity pathway. In this study, we specifically focused on CPF-induced toxicity through non-cholinergic mechanisms. Indeed, despite the well-known fact that the main property of CPF is the inhibition of acetylcholinesterase (AChE) due to its intrinsic nature, researchers have also noticed toxic effects of CPF exposure at insufficient doses to inhibit AChE, suggesting the existence of alternative so-called non-cholinergic mechanisms^66^. From the results obtained using AOP-helpFinder 2.0, a manual curation was done by experts on the identified linkages in published abstracts to avoid false positives and to retain only the most relevant articles with the significant knowledge. The highest scoring linkages (moderate, high, very high CS) were used to set up a robust foundation for the COP. AOP-helpFinder enabled the state-of-the-art of the literature concerning the stressors and events of interest. We established and enriched the COP with specialized and complementary articles to provide a comprehensive model that does not rely solely on the output of an algorithm.

Furthermore, to improve the relevance and robustness of the proposed COP, freely-accessible tools such as PubTator^67,68^ (https://www.ncbi.nlm.nih.gov/research/pubtator/) and PubPeer (https://pubpeer.com/) were used. The PubTator AI tool allows to examine which ‘bio-concepts’ (including *genes*, *diseases*, *species*, and other *chemicals*) were associated within each AI-selected article making up the COP, and PubPeer was employed for appraising the scientific authenticity of each AI-selected article, as this tool aids to detect scam articles. Moreover, articles dealing to the socio-economic impacts were also found through a classical literature search using keywords on ISI journal search engines (https://mjl.clarivate.com/home), and literature already gathered by the experts involved in this study were also added. Finally, the proposed COP was assessed in a global way using the criteria defined by the Organization for Economic and co-Operation and Development (OECD) and the AOP-Wiki guidelines^69^ to support the WoE, which is a pillar of the AOP framework.

## Data Availability

All data produced in the present work are contained in the manuscript and supplementary materials

## Acknowledgements

This work was supported by INSERM, Université Paris Cité and the French Ministries for an Ecological Transition, for Agriculture and Food, for Solidarity and Health and of Higher Education, Research and Innovation, with the financial support of the French Office for Biodiversity, as part of the call for research projects “Phytosanitary products: from exposure to impacts on human health and ecosystems”, with the fees for diffuse pollution coming from the Ecophyto II+ plan.

## Author contributions

KA conceptualization, methodology, writing-original draft & review, supervision. TC resources, methodology, data analysis, writing-original draft & review. JMB socio-economic data and critical analysis of the AO-CO link. All authors contributed to the writing and editing of the manuscript.

## Competing Interests

The authors declare they have no conflicts of interest related to this work to disclose.

## Notes

### Competing Interest Statement

The authors have declared no competing interest.

## References

1. Wild, C. P. Complementing the Genome with an “Exposome”: The Outstanding Challenge of Environmental Exposure Measurement in Molecular Epidemiology. Cancer Epidemiol. Biomarkers Prev. 14, 1847–1850 (2005).

2. Vineis, P. et al. What is new in the exposome? Environ. Int. 143, 105887 (2020).

3. Rappaport, S. M. & Smith, M. T. Environment and Disease Risks. Science 330, 460–461 (2010).

4. INSERM Collective Expertise Centre. Effects of Pesticides on Health: New Data. (EDP Sciences, Montrouge (FR), 2022).

5. Gluckman, P. D., Hanson, M. A., Cooper, C. & Thornburg, K. L. Effect of In Utero and Early-Life Conditions on Adult Health and Disease. N. Engl. J. Med. 359, 61–73 (2008).

6. Silbereis, J. C., Pochareddy, S., Zhu, Y., Li, M. & Sestan, N. The Cellular and Molecular Landscapes of the Developing Human Central Nervous System. Neuron 89, 248–268 (2016).

7. Arora, M. et al. Fetal and postnatal metal dysregulation in autism. Nat. Commun. 8, 15493 (2017).

8. Van Der Zalm, A. J. et al. A framework for establishing scientific confidence in new approach methodologies. Arch. Toxicol. 96, 2865–2879 (2022).

9. Ankley, G. T. et al. Adverse outcome pathways: A conceptual framework to support ecotoxicology research and risk assessment. Environ. Toxicol. Chem. 29, 730–741 (2010).

10. Sauer, U. G. et al. 21st Century Approaches for Evaluating Exposures, Biological Activity, and Risks of Complex Substances: Workshop highlights. Regul. Toxicol. Pharmacol. 111, 104583 (2020).

11. Kleinstreuer, N. & Hartung, T. Artificial intelligence (AI)—it’s the end of the tox as we know it (and I feel fine)*. Arch. Toxicol. (2024) doi:10.1007/s00204-023-03666-2.

12. Baker, N., Knudsen, T. & Williams, A. Abstract Sifter: a comprehensive front-end system to PubMed. F1000Research 6, Chem Inf Sci-2164 (2017).

13. Hartung, T. ToxAIcology - The evolving role of artificial intelligence in advancing toxicology and modernizing regulatory science. ALTEX 559–570 (2023) doi:10.14573/altex.2309191.

14. Lin, Z. & Chou, W.-C. Machine Learning and Artificial Intelligence in Toxicological Sciences. Toxicol. Sci. 189, 7–19 (2022).

15. Baudiffier, D., et al. Editorial trend: adverse outcome pathway (AOP) and computational strategy — towards new perspectives in ecotoxicology. Environ. Sci. Pollut. Res. 31, 6587–6596 (2023).

16. Carvaillo, J.-C., Barouki, R., Coumoul, X. & Audouze, K. Linking Bisphenol S to Adverse Outcome Pathways Using a Combined Text Mining and Systems Biology Approach. Environ. Health Perspect. 127, 047005 (2019).

17. Jaylet, T., Coustillet, T., Jornod, F., Margaritte-Jeannin, P. & Audouze, K. AOP-helpFinder 2.0: Integration of an event-event searches module. Environ. Int. 177, 108017 (2023).

18. Jornod, F. et al. AOP-helpFinder webserver: a tool for comprehensive analysis of the literature to support adverse outcome pathways development. Bioinforma. Oxf. Engl. 38, 1173–1175 (2022).

19. Jornod, F. et al. AOP4EUpest: mapping of pesticides in adverse outcome pathways using a text mining tool. Bioinformatics 36, 4379–4381 (2020).

20. Benoit, L. et al. Adverse outcome pathway from activation of the AhR to breast cancer-related death. Environ. Int. 165, 107323 (2022).

21. Jaylet, T. et al. Development of an adverse outcome pathway for radiation-induced microcephaly via expert consultation and machine learning. Int. J. Radiat. Biol. 98, 1752– 1762 (2022).

22. Jaylet, T., Quintens, R., Armant, O. & Audouze, K. An integrative systems biology strategy to support the development of adverse outcome pathways (AOPs): a case study on radiation-induced microcephaly. Front. Cell Dev. Biol. 11, 1197204 (2023).

23. Gundacker, C. et al. Reduced Birth Weight and Exposure to Per- and Polyfluoroalkyl Substances: A Review of Possible Underlying Mechanisms Using the AOP-HelpFinder. Toxics 10, 684 (2022).

24. Menzies, N. A. et al. Global burden of disease due to rifampicin-resistant tuberculosis: a mathematical modeling analysis. Nat. Commun. 14, 6182 (2023).

25. Dusza, H. M. et al. Effects of environmental pollutants on calcium release and uptake by rat cortical microsomes. NeuroToxicology 69, 266–277 (2018).

26. Nishi, T. et al. Participation of Bcl-2/Bax-α in Glutamate-induced Apoptosis of Human Glioblastoma Cells. J. Neurooncol. 44, 109–117 (1999).

27. Sunkaria, A., Sharma, D. R., Wani, W. Y. & Gill, K. D. Attenuation of Dichlorvos-Induced Microglial Activation and Neuronal Apoptosis by 4-Hydroxy TEMPO. Mol. Neurobiol. 49, 163–175 (2014).

28. Huang, L., Liu, Y., Jin, W., Ji, X. & Dong, Z. Ketamine potentiates hippocampal neurodegeneration and persistent learning and memory impairment through the PKCγ– ERK signaling pathway in the developing brain. Brain Res. 1476, 164–171 (2012).

29. Rauh, V. et al. Seven-Year Neurodevelopmental Scores and Prenatal Exposure to Chlorpyrifos, a Common Agricultural Pesticide. Environ. Health Perspect. 119, 1196– 1201 (2011).

30. Naef, N., Ciernik, A., Latal, B., Liamlahi, R., & For the Children’s Heart and Development Research Group. Hippocampal volume and cognitive performance in children with congenital heart disease. Pediatr. Res. 94, 99–102 (2023).

31. Cheong, J. L. Y. et al. Contribution of Brain Size to IQ and Educational Underperformance in Extremely Preterm Adolescents. PLoS ONE 8, e77475 (2013).

32. Bouchard, M. F. et al. Prenatal Exposure to Organophosphate Pesticides and IQ in 7-Year-Old Children. Environ. Health Perspect. 119, 1189–1195 (2011).

33. Bellanger, M., Demeneix, B., Grandjean, P., Zoeller, R. T. & Trasande, L. Neurobehavioral Deficits, Diseases, and Associated Costs of Exposure to Endocrine-Disrupting Chemicals in the European Union. J. Clin. Endocrinol. Metab. 100, 1256–1266 (2015).

34. Trasande, L. et al. Burden of disease and costs of exposure to endocrine disrupting chemicals in the E uropean U nion: an updated analysis. Andrology 4, 565–572 (2016).

35. Grosse, S. D. & Zhou, Y. Monetary Valuation of Children’s Cognitive Outcomes in Economic Evaluations from a Societal Perspective: A Review. Children 8, 352 (2021).

36. Nevin, R. How Lead Exposure Relates to Temporal Changes in IQ, Violent Crime, and Unwed Pregnancy. Environ. Res. 83, 1–22 (2000).

37. Reyes, J. W. Environmental Policy as Social Policy? The Impact of Childhood Lead Exposure on Crime. BE J. Econ. Anal. Policy 7, (2007).

38. Gould, E. Childhood Lead Poisoning: Conservative Estimates of the Social and Economic Benefits of Lead Hazard Control. Environ. Health Perspect. 117, 1162–1167 (2009).

39. Lin, D., Lutter, R. & Ruhm, C. J. Cognitive performance and labour market outcomes. Labour Econ. 51, 121–135 (2018).

40. Pichery, C. et al. Childhood lead exposure in France: benefit estimation and partial cost-benefit analysis of lead hazard control. Environ. Health 10, 44 (2011).

41. Olesen, J. et al. The economic cost of brain disorders in Europe. Eur. J. Neurol. 19, 155–162 (2012).

42. Organisation for Economic Co-operation and Development. Valuing the Avoidance of IQ Losses in Children: A Large Scale Multi-Country Stated Preference Approach. vol. 219 https://www.oecd-ilibrary.org/environment/valuing-the-avoidance-of-iq-losses-in-children_71574eb4-en (2023).

43. Soguel, N. & Griethuysen, P. V. Cost of Illness and Contingent Valuation : Controlling for the Motivations of Expressed Preferences in an Attempt to Avoid Double-Counting. Économie PubliquePublic Econ. (2004) doi:10.4000/economiepublique.400.

44. Quilez-Robres, A., González-Andrade, A., Ortega, Z. & Santiago-Ramajo, S. Intelligence quotient, short-term memory and study habits as academic achievement predictors of elementary school: A follow-up study. Stud. Educ. Eval. 70, 101020 (2021).

45. Liu, J., Hwang, W.-T., Dickerman, B. & Compher, C. Regular breakfast consumption is associated with increased IQ in kindergarten children. Early Hum. Dev. 89, 257–262 (2013).

46. Yang, S. et al. Associations of screen use with cognitive development in early childhood: the ELFE birth cohort. J. Child Psychol. Psychiatry jcpp.13887 (2023) doi:10.1111/jcpp.13887.

47. Carloni, M. et al. The impact of early life permethrin exposure on development of neurodegeneration in adulthood. Exp. Gerontol. 47, 60–66 (2012).

48. Vorstman, J. A. S. et al. Autism genetics: opportunities and challenges for clinical translation. Nat. Rev. Genet. 18, 362–376 (2017).

49. Canitano, R. & Palumbi, R. Excitation/Inhibition Modulators in Autism Spectrum Disorder: Current Clinical Research. Front. Neurosci. 15, 753274 (2021).

50. Taylor, D. L., Jones, F., Kubota, E. S. F. C. S. & Pocock, J. M. Stimulation of Microglial Metabotropic Glutamate Receptor mGlu2 Triggers Tumor Necrosis Factor α-Induced Neurotoxicity in Concert with Microglial-Derived Fas Ligand. J. Neurosci. 25, 2952–2964 (2005).

51. Slikker, W. Cognitive Tests: Interpretation for Neurotoxicity? (Workshop Summary). Toxicol. Sci. 58, 222–234 (2000).

52. European Chemicals Agency. Lead in shot, bullets and fishing weights. (2022).

53. European Chemicals Agency. ECHA proposes a restriction on lead compounds in PVC articles. (2017).

54. French High Council for Public Health. Determination of new lead exposure risk management objectives. (2014).

55. Teeguarden, J. G. et al. Completing the Link between Exposure Science and Toxicology for Improved Environmental Health Decision Making: The Aggregate Exposure Pathway Framework. Environ. Sci. Technol. 50, 4579–4586 (2016).

56. Hines, D. E., Edwards, S. W., Conolly, R. B. & Jarabek, A. M. A Case Study Application of the Aggregate Exposure Pathway (AEP) and Adverse Outcome Pathway (AOP) Frameworks to Facilitate the Integration of Human Health and Ecological End Points for Cumulative Risk Assessment (CRA). Environ. Sci. Technol. 52, 839–849 (2018).

57. Luengo-Fernandez, R., Leal, J., Gray, A. & Sullivan, R. Economic burden of cancer across the European Union: a population-based cost analysis. Lancet Oncol. 14, 1165–1174 (2013).

58. Wang, Y. C., McPherson, K., Marsh, T., Gortmaker, S. L. & Brown, M. Health and economic burden of the projected obesity trends in the USA and the UK. The Lancet 378, 815–825 (2011).

59. Wang, H. et al. Modelling the economic burden of SARS-CoV-2 infection in health care workers in four countries. Nat. Commun. 14, 2791 (2023).

60. Dereumeaux, C. et al. Biomarkers of exposure to environmental contaminants in French pregnant women from the Elfe cohort in 2011. Environ. Int. 97, 56–67 (2016).

61. Béranger, R. et al. Multiple pesticide analysis in hair samples of pregnant French women: Results from the ELFE national birth cohort. Environ. Int. 120, 43–53 (2018).

62. Tagne-Fotso, R., et al. Exposure of the General French Population to Herbicides, Pyrethroids, Organophosphates, Organochlorines, and Carbamate Pesticides in 2014–2016: Results from the Esteban Study. https://www.ssrn.com/abstract=4452103 (2023) doi:10.2139/ssrn.4452103.

63. French Agency for Food, Environmental and Occupational Health & Safety (ANSES). French Total Diet Study 2: Pesticide Residues, Additives, Acrylamide, Polycyclic Aromatic Hydrocarbons. https://www.anses.fr/fr/system/files/PASER2006sa0361Ra2.pdf (2011).

64. French Agency for Food, Environmental and Occupational Health & Safety (ANSES). Pesti’home Study: National Survey of Domestic Pesticide Use. https://www.anses.fr/fr/system/files/2019Pestihome.pdf (2019).

65. Auburtin, G., Lecomte, J. & Moreau, J. L’utilisation Des Biocides En Milieu Domestique et La Perception Des Risques Liés à Cette Utilisation Dans Une Population Française. https://primequal.fr/sites/default/files/auburtint_rf.pdf (2005).

66. Van Melis, L. V. J., Heusinkveld, H. J., Langendoen, C., Peters, A. & Westerink, R. H. S. Organophosphate insecticides disturb neuronal network development and function via non-AChE mediated mechanisms. NeuroToxicology 94, 35–45 (2023).

67. Wei, C.-H., Kao, H.-Y. & Lu, Z. PubTator: a web-based text mining tool for assisting biocuration. Nucleic Acids Res. 41, W518–W522 (2013).

68. Wei, C.-H., Allot, A., Leaman, R. & Lu, Z. PubTator central: automated concept annotation for biomedical full text articles. Nucleic Acids Res. 47, W587–W593 (2019).

69. Organisation for Economic Co-operation and Development. Users’ Handbook Supplement to the Guidance Document for Developing and Assessing Adverse Outcome Pathways. vol. 1 https://www.oecd-ilibrary.org/environment/users-handbook-supplement-to-the-guidance-document-for-developing-and-assessing-adverse-outcome-pathways_5jlv1m9d1g32-en (2018).

