## supplementary for "The Cost Outcome Pathway Framework: Integrating socio-economic impacts to Adverse Outcome Pathways"

### Table of Contents

#### Supplementary Tables

**Supplementary Table 1. Full distribution of stressor-event relationships from AOP-helpFinder results**. For convenience, two sheets are supplied. Sheet 'stressor-event' provides an overview of stressor-based links, while Sheet 'event-stressor' provides an overview of event-based links. Data between the two sheets is strictly identical.
https://github.com/systox1124/bioRxiv-COP-publication.git

**Supplementary Table 2. Full distribution of event-event relationships from AOP-helpFinder results**.
https://github.com/systox1124/bioRxiv-COP-publication.git

**Supplementary Table 3. PubTator bioconcepts found for each article used in the development of the COP**. Sheet 'all_bioconcepts' includes all the bio-concepts detected in each category for each article. Sheet 'top_bioconcepts' contains only the most frequently detected bio-concept for each category for each article.
https://github.com/systox1124/bioRxiv-COP-publication.git

**Supplementary Table 4: List of the 24 pesticides.** The selected chemicals were divided into two main categories: organophosphates (n=3) and their metabolites (n=4), and pyrethroids of type I (n=7), of type II (n=6) and their metabolites (n=3). Piperonyl-butoxide was considered as a separate category. a, Dereumeaux et al., 2016; b, Béranger et al., 2020; c, Tagne-Fosto et al., 2022; d, Anses, 2011; e, Cnam_IHIE, 2005; f, Anses, 2019; g, BNVD, 2011-2015. 3-BPA, 3-phenoxybenzoic acid; Cl2CA , cis-3-(2,2dichlorovinyl)-2,2-dimethylcyclopropanecarboxylic acid; ClCF3CA, 3-(2-chloro-3,3,3-trifluoro-1-propenyl)-2,2-dimethylcyclopropanecarboxylic acid; TCPy, 3,5,6-trichloro-2-pyridinol.

**Supplementary Table 5. List of the 248 conventional events (MIE, KE, AO) and five CO-related terms used for searches with AOP-helpFinder v2**.
https://github.com/systox1124/bioRxiv-COP-publication.git

### Supplementary Figures

**Supplementary Figure 1.** Distribution of stressor-event links according to the 23 most common stressors, representing 100% of the total dataset (23 stressors and 2404 links). No link was found for the stressor CICF3CA.

**Supplementary Figure 2.** Distribution of stressor-event links according to the 20 most common events, representing 74.4% of the total dataset (141 distinct events and 2404 links).

**Supplementary Figure 3.** Distribution of event-event links according to the 20 most common events, representing 61.0% of the total dataset (225 distinct events and 73079 links).

### KER Justification

Each KER's argumentation.

### Supplementary references

**Supplementary Table 4: List of the 24 pesticides.** The selected chemicals were divided into two main categories: organophosphates (n=3) and their metabolites (n=4), and pyrethroids of type I (n=7), of type II (n=6) and their metabolites (n=3). Piperonyl-butoxide was considered as a separate category. a, Dereumeaux et al., 2016[1]; b, Béranger et al., 2020[2]; c, Tagne-Fosto et al., 2022[3]; d, Anses, 2011[4]; e, Cnam_IHIE, 2005[5]; f, Anses, 2019[6]; g, BNVD, 2011-2015. 3-BPA, 3-phenoxybenzoic acid; Cl2CA , cis-3-(2,2dichlorovinyl)-2,2-dimethylcyclopropane-carboxylic acid; ClCF3CA, 3-(2-chloro-3,3,3-trifluoro-1-propenyl)-2,2-dimethylcyclopropanecarboxylic acid; TCPy, 3,5,6-trichloro-2-pyridinol.

| **Family** | **Substance name** | **CAS number** | **Type** | **French HBM data** | | | **Food contamination in France** | **Domestic use in France** | | **Agricultural use in France 2011-2015** |
|---|---|---|---|---|---|---|---|---|---|---|
| | | | | **Elfe cohort (detected in urine)[a]** | **Elfe cohort (detected in hair)[b]** | **Esteban study (detected in urine)[c]** | **EAT2 (detected)[d]** | **Cnam-IHIE study[e]** | **Pesti'HOME study[f]** | **BnVD database[g]** |
| Pyrethroid | **Allethrin** | 584-79-2 | type I pyrethroid | | | | | x | | x |
| | **Bifenthrin** | 82657-04-3 | type I pyrethroid | | | | x | x | x | x |
| | **Cyfluthrin** | 68359-37-5 | type II pyrethroid | | | | x | x | x | x |
| | **Cypermethrin** | 52315-07-8 | tenfantype II pyrethroid | | x | | | x | x | x |
| | **Deltamethrin** | 52918-63-5 | type II pyrethroid | | | | | x | x | x |
| | **lambda-cyhalothrin** | 91465-08-6 | type II pyrethroid | | x | | x | | x | x |
| | **Esfenvalerate** | 66230-04-4 | type II pyrethroid | | | | | | x | x |
| | **Permethri** | 52645- | type I | | x | | x | x | x | x |

| | | | | | | | | | | |
|---|---|---|---|---|---|---|---|---|---|---|
| | **n** | 53-1 | pyrethroid | | | | | | | |
| | **Resmethrin** | 10453-86-8 | type I pyrethroid | | | | | | x | x |
| | **Phenothrin** | 26002-80-2 | type I pyrethroid | | | | | x | x | |
| | **tau-fluvalinate** | 102851-06-9 | type II pyrethroid | | | | | | x | x |
| | **Tefluthrin** | 79538-32-2 | type I pyrethroid | | | | | | | x |
| | **Tetramethrin** | 7696-12-0 | type I pyrethroid | | | | | x | | x |
| | **(3-PBA)** | 3739-38-6 | metabolite (various pyrethroids) | x | x | | | | | |
| | **Cl2CA** | 55701-05-8 | metabolite (cyflutrhin) | x | x | | | | | |
| | **ClCF3CA** | 72748-35-7 | metabolite (teflutrhin) | | x | | | | | |
| Organophosphate | **Chlorpyrifos** | 2921-88-2 | organophosphate | | | x | x | x | x | x |
| | **Chlorpyrifos-methyl** | 5598-13-0 | organophosphate | | | x | x | | | x |
| | **Ethoprophos** | 13194-48-4 | organophosphate | | | | x | | | x |
| | **Chlorpyrifos-oxon** | 5598-15-2 | metabolite (chlorpyrifos) | | | x | | | | |
| | **TCPy** | 6515-38-4 | metabolite (chlorpyrifos) | | x | x | | | | |
| | **diethyl-** | 598-02-7 | metab | x | x | x | | | | |

| | | | | | | | | | | |
|---|---|---|---|---|---|---|---|---|---|---|
| | **phosphate** | | olite (various organophosphates) | | | | | | | |
| | **diethyl-thiophosphate** | 2465-65-8 | metabolite (various organophosphates) | **x** | **x** | **x** | | | | |
| Synergist | **Piperonyl butoxide** | 51-03-6 | pyrethroid synergist | | | | **x** | | **x** | **x** |

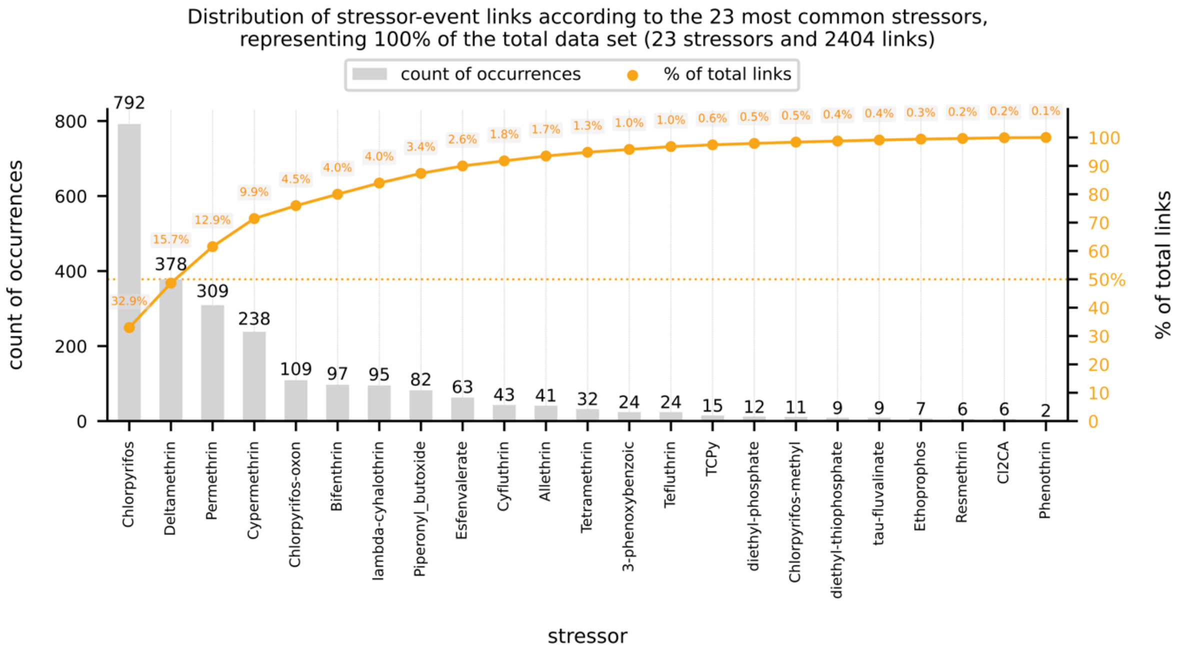


**Supplementary Figure 1.** Distribution of the identified stressor-event linkage in the literature using AOP-helpFinder for the 23 retrieved stressors, representing 100% of the total dataset (23 stressors and 2404 links). No link was found for the pyrethroid metabolite of tefluthrin (ClCF3CA).

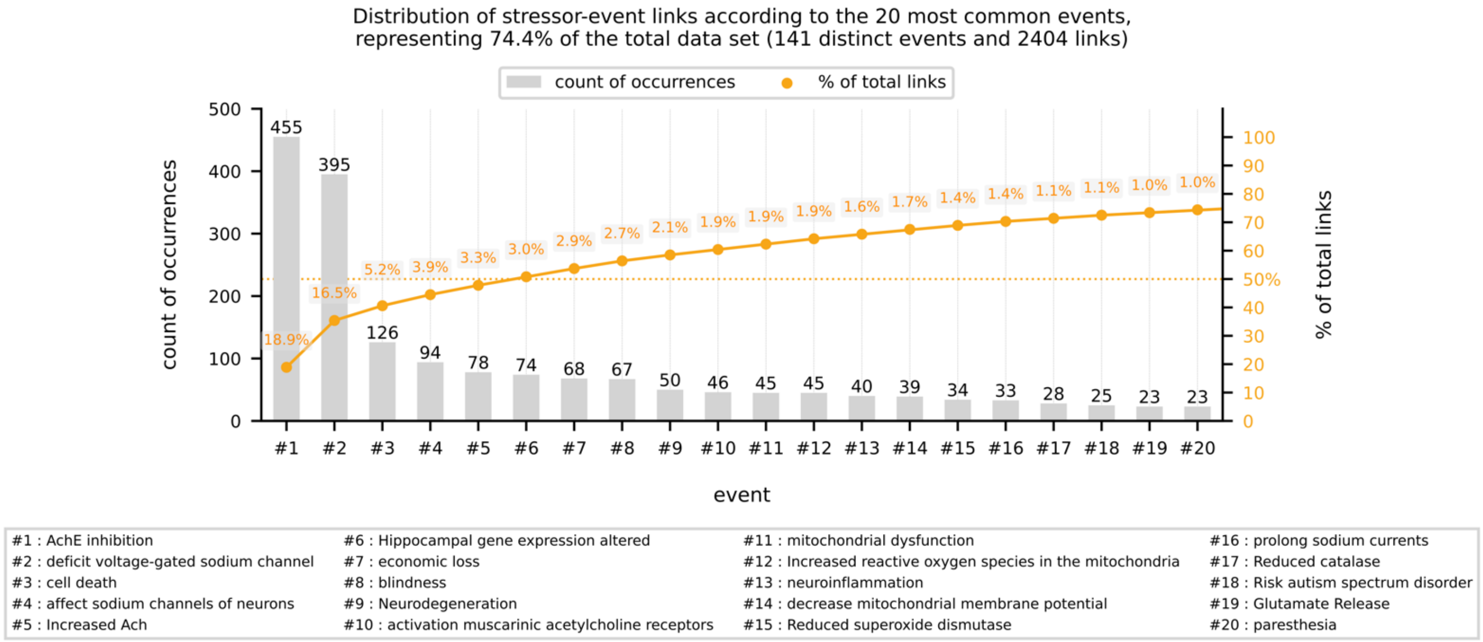


**Supplementary Figure 2.** Distribution of stressor-event links according to the 20 most common events, representing 74.4% of the total dataset (141 distinct events and 2404 links).

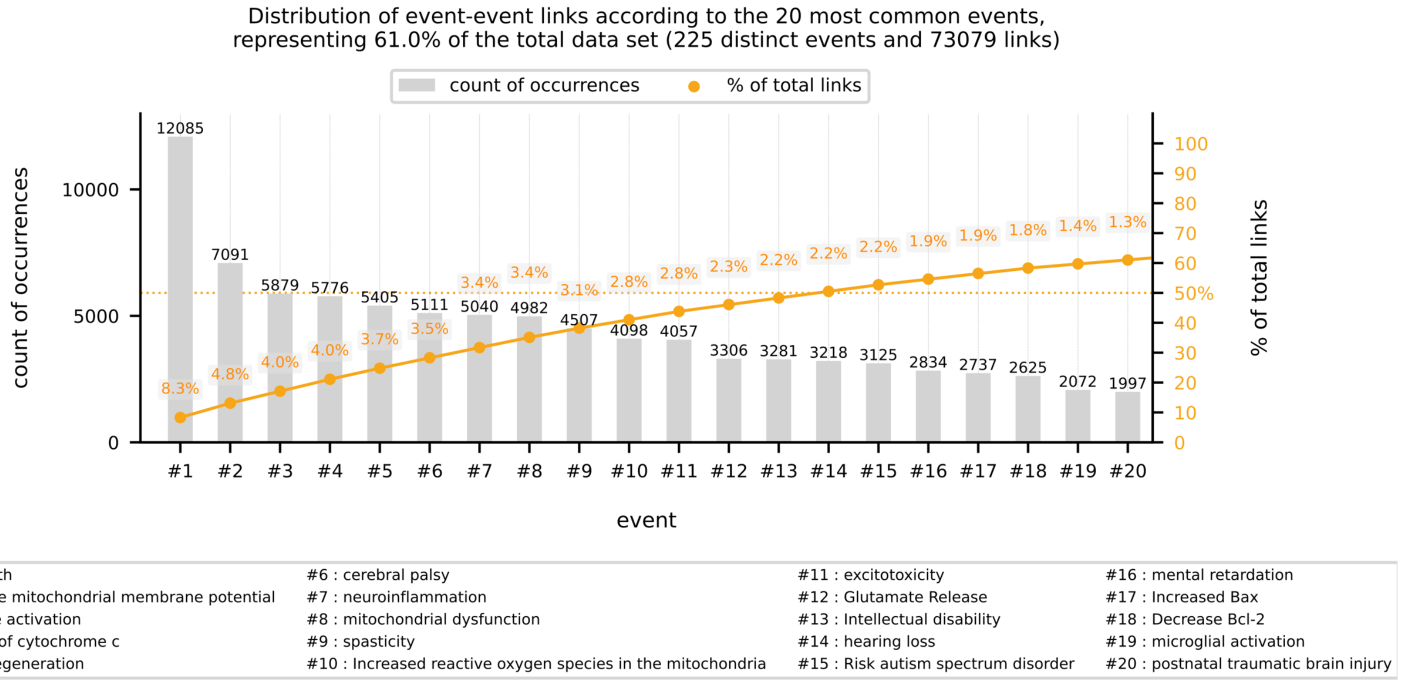


**Supplementary Figure 3.** Distribution of event-event links according to the 20 most common events, representing 61.0% of the total dataset (225 distinct events and 73079 links).

**KER justification:**

1- **KER 1-2: Activation, IP3R (KE ID 2037) and Activation, RyR (KE ID 2186) lead to Increased, glutamate (KE ID 1350*).***

IP3R and RyRs are expressed by many types of neurons[7]. Once they are activated (e.g., OPs such as CPF or TOCP (tri-o-cresyl phosphate)), a release of $Ca^{2+}$ from intracellular stores occurs in the cytoplasm which triggers the exocytosis of glutamate. Separately, IP3Rs and RyRs agonists were capable of increasing glutamate levels in nerve tissue, while their antagonists did not reverse the effect, suggesting the participation of both receptors in the glutamate homeostasis[8,9].

2- **KER 3-4-9-10: Increased, glutamate (KE ID 1350) leads to decreased, Bcl-2 expression (KE ID 2123) and increased, Bax expression (KE ID 2124), resulting in cell injury/death (KE ID 55)**

Glutamate is a major excitatory neurotransmitter in the mammalian central nervous system (CNS). Neurons and glial cells, including microglia, express glutamate receptors. Bax activation leads to the mitochondrial release of cytochrome c leading to the activation of caspases (3 & 9) and subsequently to apoptosis[10]. In several CNS cell lines[11,12], glutamate down-regulates Bcl-2 with a concomitant up-regulation of Bax. Moreover, Lee *et al.* showed that CPF increases the activity of caspases 3 and 9 in human embryonic stem cell-derived neural precursor cells (hNPCs) and cytochrome c mitochondrial release and cell death[13].

3- **KER 5-6-7-8: Increased, glutamate (KE ID 1350) leads to Activation, microglia (KE ID 1998) and Activation, microglia leads to neuroinflammation (KE ID 188), resulting in cell injury/death (KE ID 55)**

Neuroinflammation largely depends on microglia which are resident immune cells of the nerve tissue, constantly scavenging the CNS for damaged neurons, foreign material, or infectious agents (mainly through phagocytosis). In addition to releasing pro-inflammatory mediators such as IL-1β or TNF-α, microglial cells could also deliver cytotoxic effectors such as proteases, reactive oxygen intermediates and nitrite oxide (NO)[14] (immunoexcitotoxicity). Glutamate-induced overstimulation of microglia leads to microglia-induced neuroinflammation, which in turn can strengthen inflammation by operating in an autocrine fashion[15]. Dichlorvos (an OP) exposure activates microglia and causes secretion of IL-1β, TNF-α, and NO leading to neuronal death[16] *via* p53, Bax, cytochrome c and caspase 3. In summary, a continuous neuroinflammation (due to the persistence of stressors) through the release of pro-inflammatory cytokines (IL-1β, TNF-α) and NO, triggers apoptotic processes (involving p53, Bax, cytochrome c and caspase 3, for example) which ultimately leads to cell death (immunoexcitotoxicity).

### *4-* KER 11: cell injury/death (KE ID 55) leads to Hippocampal Physiology, Altered (KE ID 758) resulting in Impairment, learning and memory (KE ID 341).

Stresses during childhood can lead to lowered hippocampal volume and neurogenesis in humans and animals[17] due to impaired maturation of the brain. Indeed, in adolescent rats (PND 28-48, a window of vulnerability), alcohol exposure results in a reduced neurogenesis with lowered differentiating neurons that were due at least to an increase of cell death of immature neurons rather than a decrease in neural progenitor cell proliferation[18]. Interestingly, the loss of neurogenesis could be both period- and region- specific. In another study, cell death in the CA1 region (of the Ammon's corn) and in the dentate gyrus in PND-11 Sprague–Dawley rats leads to a decline in memory and learning efficiency later in life (PND-60, young adults); indeed, these rats showed poorer results in the Morris water maze test, both during the training and testing stages[19]. Difficulties in the training phase reflect learning disorders, while difficulties in the testing phase reflect memory disorders. Besides, authors showed that the hippocampal neurodegeneration could partly occur through Bcl-2 downregulation. On the other hand, other researchers observed longer times and distances to reach the platform in the Morris water maze test in rats with a 2.3% reduction in hippocampal volume[20].

**Supplementary references**


1. Dereumeaux, C. *et al.* Biomarkers of exposure to environmental contaminants in French pregnant women from the Elfe cohort in 2011. *Environ. Int.* **97**, 56–67 (2016).
2. Béranger, R. *et al.* Multiple pesticide analysis in hair samples of pregnant French women: Results from the ELFE national birth cohort. *Environ. Int.* **120**, 43–53 (2018).
3. Tagne-Fotso, R. *et al. Exposure of the General French Population to Herbicides, Pyrethroids, Organophosphates, Organochlorines, and Carbamate Pesticides in 2014–2016: Results from the Esteban Study*. https://www.ssrn.com/abstract=4452103 (2023) doi:10.2139/ssrn.4452103.
4. French Agency for Food, Environmental and Occupational Health & Safety (ANSES). *French Total Diet Study 2: Pesticide Residues, Additives, Acrylamide, Polycyclic Aromatic Hydrocarbons*. https://www.anses.fr/fr/system/files/PASER2006sa0361Ra2.pdf (2011).
5. Auburtin, G., Lecomte, J. & Moreau, J. *L'utilisation Des Biocides En Milieu Domestique et La Perception Des Risques Liés à Cette Utilisation Dans Une Population Française*. https://primequal.fr/sites/default/files/auburtint_rf.pdf (2005).
6. French Agency for Food, Environmental and Occupational Health & Safety (ANSES). *Pesti'home Study: National Survey of Domestic Pesticide Use*. https://www.anses.fr/fr/system/files/2019Pestihome.pdf (2019).
7. Ogawa Y. & Murayama T. Ryanodine receptors in the central nervous system. *Folia Pharmacol. Jpn.* **105**, 423–430 (1995).
8. Yamamura, S. *et al.* Effects of zonisamide on neurotransmitter release associated with inositol triphosphate receptors. *Neurosci. Lett.* **454**, 91–96 (2009).
9. Mori, F., Okada, M., Tomiyama, M., Kaneko, S. & Wakabayashi, K. Effects of ryanodine receptor activation on neurotransmitter release and neuronal cell death following kainic acid-induced status epilepticus. *Epilepsy Res.* **65**, 59–70 (2005).
10. Zou, H., Li, Y., Liu, X. & Wang, X. An APAF-1.cytochrome c multimeric complex is a functional apoptosome that activates procaspase-9. *J. Biol. Chem.* **274**, 11549–11556 (1999).
11. Nishi, T. *et al.* Participation of Bcl-2/Bax-α in Glutamate-induced Apoptosis of Human Glioblastoma Cells. *J. Neurooncol.* **44**, 109–117 (1999).
12. Gao, M. *et al.* Pinocembrin prevents glutamate-induced apoptosis in SH-SY5Y neuronal cells via decrease of bax/bcl-2 ratio. *Eur. J. Pharmacol.* **591**, 73–79 (2008).
13. Lee, J. E., Lim, M. S., Park, J. H., Park, C. H. & Koh, H. C. Nuclear NF-κB contributes to chlorpyrifos-induced apoptosis through p53 signaling in human neural precursor cells. *NeuroToxicology* **42**, 58–70 (2014).
14. Gehrmann, J., Matsumoto, Y. & Kreutzberg, G. W. Microglia: intrinsic immuneffector cell of the brain. *Brain Res. Brain Res. Rev.* **20**, 269–287 (1995).
15. Iovino, L., Tremblay, M. E. & Civiero, L. Glutamate-induced excitotoxicity in Parkinson's disease: The role of glial cells. *J. Pharmacol. Sci.* **144**, 151–164 (2020).
16. Sunkaria, A., Sharma, D. R., Wani, W. Y. & Gill, K. D. Attenuation of Dichlorvos-Induced Microglial Activation and Neuronal Apoptosis by 4-Hydroxy TEMPO. *Mol. Neurobiol.* **49**, 163–175 (2014).
17. Malave, L., Van Dijk, M. T. & Anacker, C. Early life adversity shapes neural circuit function during sensitive postnatal developmental periods. *Transl. Psychiatry* **12**, 306 (2022).
18. Broadwater, M. A., Liu, W., Crews, F. T. & Spear, L. P. Persistent Loss of Hippocampal Neurogenesis and Increased Cell Death following Adolescent, but Not Adult, Chronic Ethanol Exposure. *Dev. Neurosci.* **36**, 297–305 (2014).

19. Huang, L., Liu, Y., Jin, W., Ji, X. & Dong, Z. Ketamine potentiates hippocampal neurodegeneration and persistent learning and memory impairment through the PKCγ–ERK signaling pathway in the developing brain. *Brain Res.* **1476**, 164–171 (2012).
20. Su, J., Sripanidkulchai, K., Hu, Y., Wyss, J. M. & Sripanidkulchai, B. The Effect of Ovariectomy on Learning and Memory and Relationship to Changes in Brain Volume and Neuronal Density. *Int. J. Neurosci.* **122**, 549–559 (2012).
